# The ventral striatum harbours patient specific intracranial neural signatures of obsessions and compulsions

**DOI:** 10.1101/2021.04.04.21254830

**Authors:** Egill A Fridgeirsson, MN Bais, N Eijsker, RM Thomas, DJA Smit, IO Bergfeld, PR Schuurman, P van den Munckhof, P de Koning, N Vulink, M Figee, A Mazaheri, GA van Wingen, D Denys

## Abstract

Deep brain stimulation is a treatment option for patients with refractory obsessive-compulsive disorder. A new generation of stimulators hold promise for closed loop stimulation, with adaptive stimulation in response to biological signals Here we aimed to discover a suitable biomarker in the ventral striatum in patients with obsessive compulsive disorder using local field potentials. We induced obsessions and compulsions and trained a deep learning model on the recorded time series. Average classification sensitivities were 47% for obsessions and 66% for compulsions for patient specific models at 25% chance level. Sensitivity for obsessions reached over 90% in one patient, whereas performance was near chance level when the model was trained across patients. Optimal sensitivity for obsessions and compulsions was obtained at different recording sites. This study shows that closed loop stimulation is a viable option for OCD, but that intracranial biomarkers for obsessive-compulsive disorder are patient and not disorder specific.

## Introduction

Obsessive compulsive disorder (OCD) is a psychiatric disorder characterized by intrusive obsessive thoughts (obsessions) and repetitive behaviors (compulsions). Approximately 10% of OCD patients does not respond and continues to experience symptoms despite routine treatment with serotonin reuptake inhibitors, antipsychotics and behavioral therapy ^1^. For those severe treatment resistant patients, deep brain stimulation (DBS) is an emerging treatment which has around 50-60% responder rate ^2–5^. In this neurosurgical treatment electrodes are implanted in specific brain regions which can then be stimulated with electrical pulses. For OCD, electrodes are most often implanted in striatal regions such as nucleus accumbens (NAc), ventral capsules/ventral striatum (VC/VS) and ventral anterior limb of the internal capsules (vALIC)^6,7^.

Typically, DBS delivers the electrical pulses continuously at a constant level, as opposed to closed loop stimulation where the target region is stimulated in response to some aberrant neural activity. Continuous stimulation requires time consuming iterative adjustments of stimulation parameters until maximal treatment response is achieved, while closed loop stimulation, once a suitable biomarker has been identified, could adapt its stimulation response automatically to that biomarker. This requires a device that can both stimulate and record neural activity, as well as be programmable with models to make the stimulation react to the neural activity. Devices that can both stimulate and sense have in the last years become available such as the Activa PC+S prototype ^8^ and the recently commercially available Percept PC^a^, both from Medtronic.

Closed loop stimulation has been explored in movement disorders such as essential tremor and Parkinson’s disease ^9^, with some candidate biomarkers identified^10,11^. Two studies on essential tremor used machine learning algorithms to control closed loop stimulation in a small sample of patients ^12,13^. They show that it is possible to perform closed loop stimulation that is equivalent to continuous stimulation while reducing energy needs. On the other hand efforts for developing closed loop stimulation for OCD are still at the relatively early stage of finding the most suitable biomarker ^14^. So far, only two case-studies in OCD using chronic LFP recordings from PC+S have been published. In the first study, data from a patient with STN leads^15^ showed theta oscillations in the ventral STN that were not found in the STN recordings from a comparison group of PD patients. Theta oscillations decreased with OCD symptom provocation and correlated inversely with OCD severity over time. In the second case-study, a PC+S device was used to simultaneously record and stimulate in the supplementary motor area (SMA) and ventral capsule/ventral striatum (VC/VS) in a patient with OCD.^16^ They used frequency mismatched stimulation and hypothesized that such stimulation would reduce compulsive symptoms. While the patient reported subjective improvement, no such improvement was seen in the clinical rating scales.

To date there has not yet been a clear biomarker to be used for closed loop stimulation in OCD. Here we have a unique dataset of 11 patients with consistent LFP recordings during symptom provocation. We investigated whether a candidate biomarker could be derived using deep learning on minimally processed raw time-series data recorded from the DBS electrodes during symptom provocation. Deep learning has transformed whole domains such as natural language processing and computer vision ^17^ and in general has shown promise where it is used on structured data. For our goal of time-series classification, a deep learning model has previously achieved state of the art results ^18^.

In order to elicit OCD symptoms and the accompanying neural state, we used a symptom provocation paradigm that was inspired by the induction of symptoms during exposure therapy. We then tested if we could predict the symptomatic state from baseline using the brain recordings from the DBS leads implanted in the vALIC using the investigational Activa PC+S from Medtronic. We further explored whether a biomarker would be generic across OCD patients or whether such biomarkers would be patient specific. Therefore, we tested if one model can be built on the group-level to predict symptom states in a new patient, and if a patient specific model could predict symptom states in the same patient at later time

## Results

### Symptom provocation

Eleven patients (10 women, age range 35-68) with refractory OCD underwent surgery for bilateral placement of DBS leads in the vALIC (Table 1). The leads were placed such that the most ventral contact point was in the NAc in the plane 3mm anterior to the anterior commissure (see supplemental Figure 1). Two to three weeks after, before DBS was turned on, patients performed a symptom provocation task while LFPs were recorded. First the patient was asked to sit calmly and watch a movie. Then obsessions were induced in a patient specific manner, for example by asking the patient to touch the floor (Table 2). In the third phase the patient was allowed to act out their compulsions, for example by washing their hands. Lastly when the patient was relieved of her obsessions and compulsions, they were instructed to sit again calmly. Before and after each task, patients rated their symptoms on visual analog scales (VAS) across six different dimensions (Figure 2a). These tasks were repeated for four rounds.

**Table 1:**
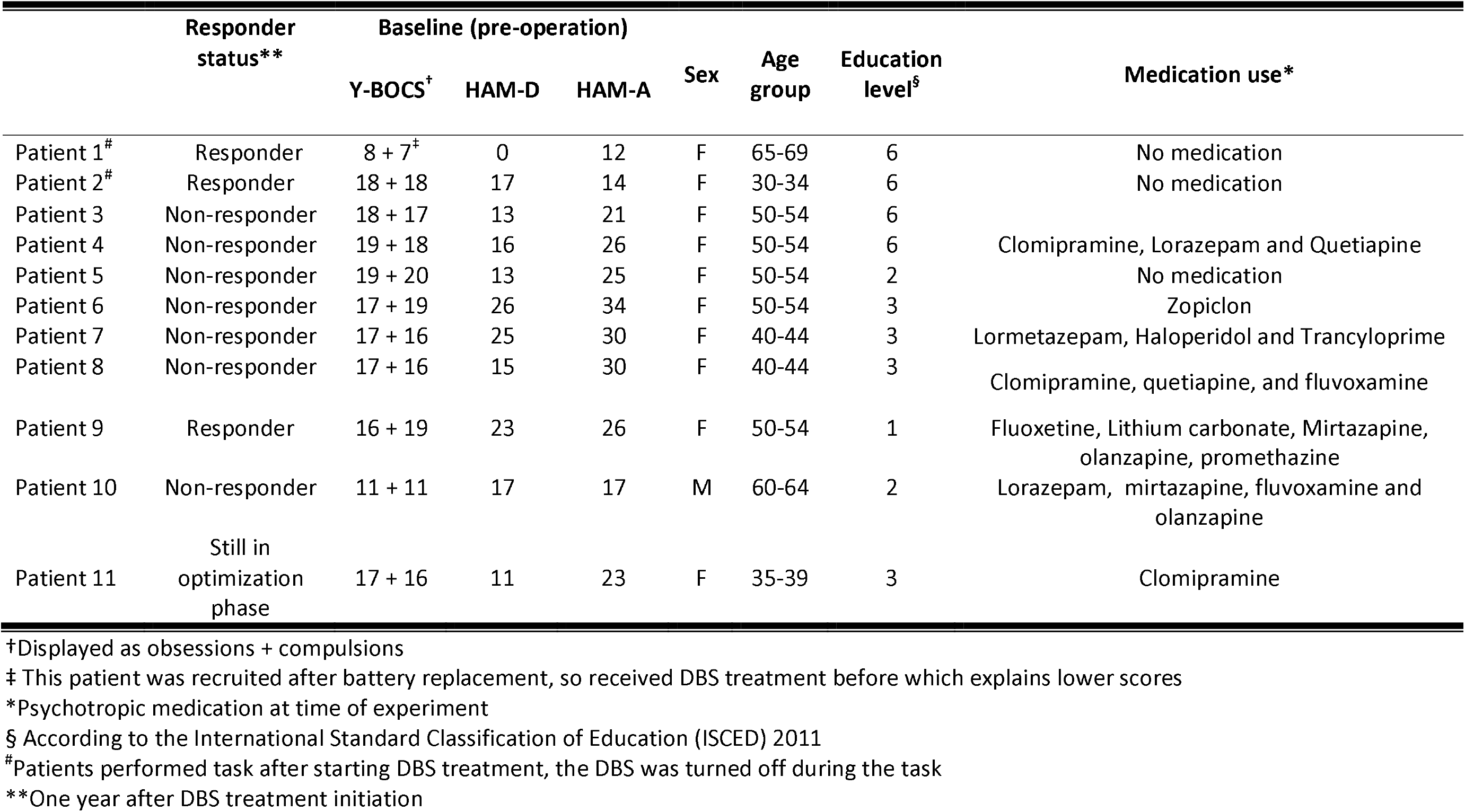
Patient demographics

**Table 2:** Symptom provocations

|  | Subtype | Symptom provocation | Compulsive behavior |
| --- | --- | --- | --- |
| Patient 1 | Contamination | Touching the floor | Washing hands |
| Patient 2 | Religious | Reading cards with religious texts | Praying, writing |
| Patient 3 | Contamination / Checking | Touch floor near toilet | Washing hands |
| Patient 4 | Contamination / Checking | Other people touching a mug from home | Washing/checking the mug |
| Patient 5 | Contamination/Symmetry | Touch the floor | Washing hands |
| Patient 6 | Harm / Symmetry | Hair dryer plugged in in another room | Check hair dryer, unplug it. |
| Patient 7 | Contamination / Symmetry | Touch the floor | Washing hands |
| Patient 8 | Contamination | Touch doorknob/light switch | Washing hands |
| Patient 9 | Harm / Contamination | Think about incident involving a contaminated mattress at home | Look at pictures of mattress to check if it's clean |
| Patient 10 | Religious | Reading the bible/ looking at pictures from unsettling passages | Read reassuring passages |
| Patient 11 | Contamination | Touching the floor | Washing hands |

**Figure 2:**
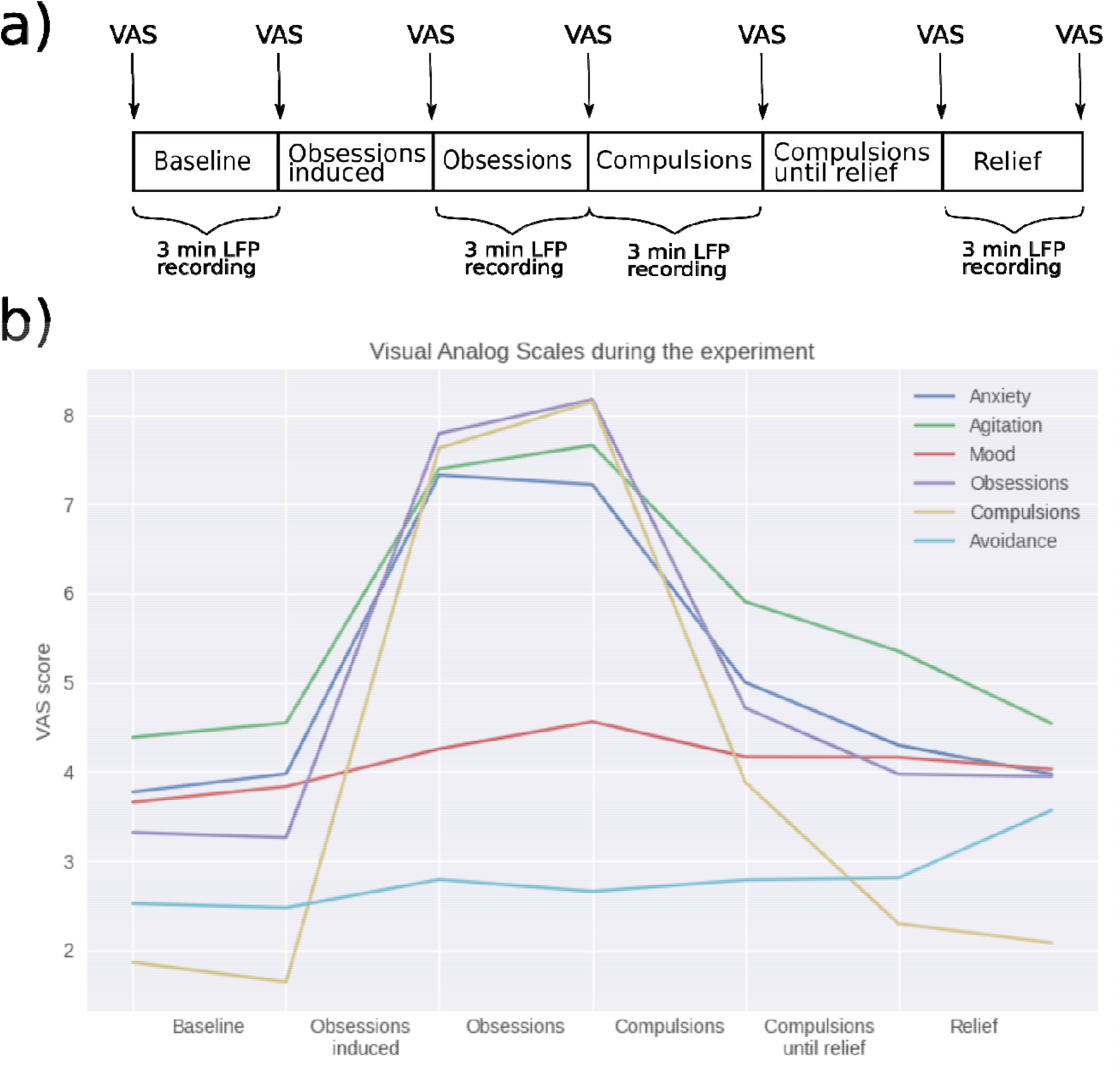
a) Timeline of experiment and VAS measurements b) Visual analog scores averaged over patients during task performance.

**Figure 3:**
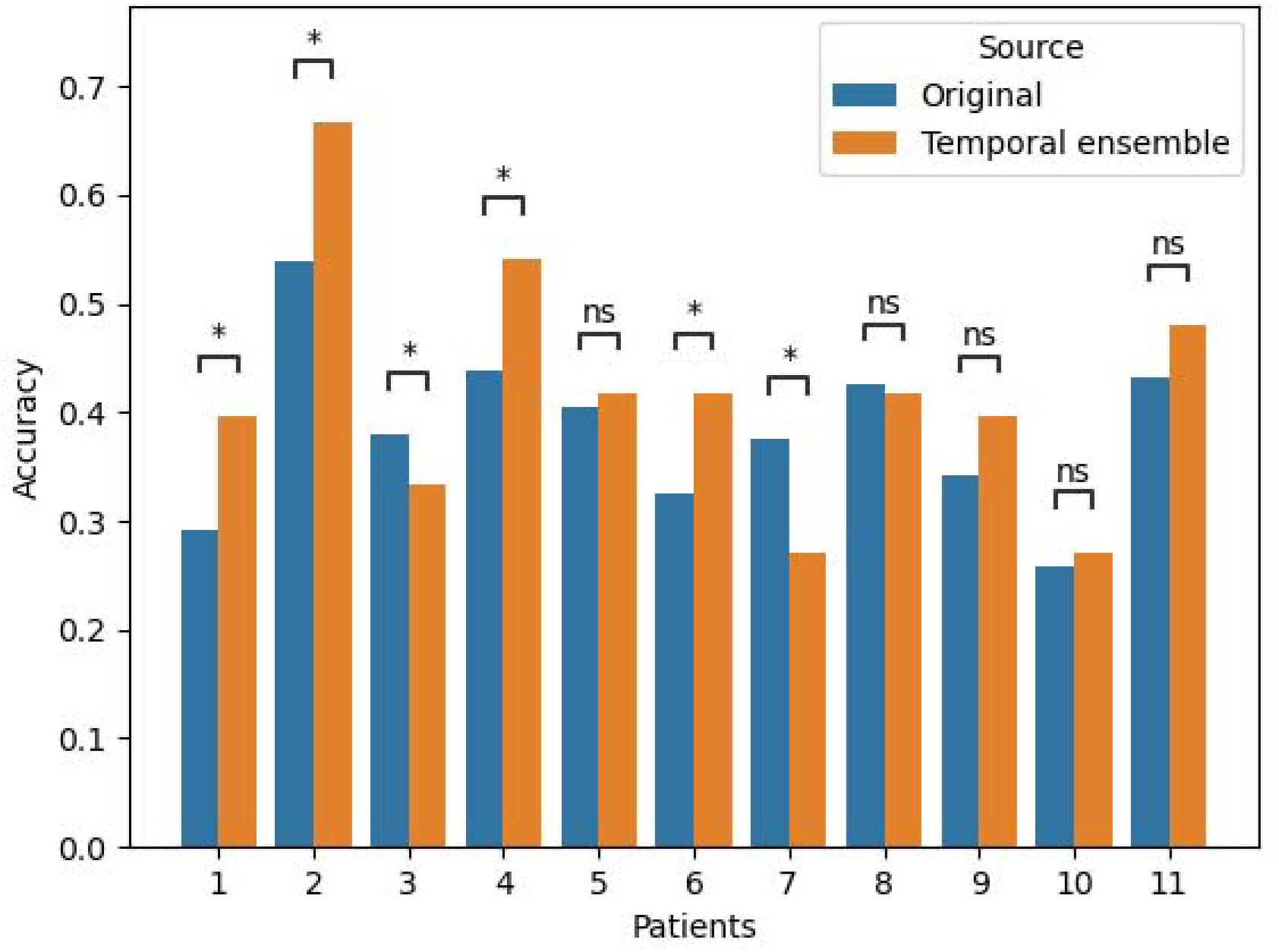
Effect of temporal ensemble on the accuracy. *p<0.05, ns = not significant.

As can be seen from Figure 2b all the measured VAS scores except mood and avoidance rose sharply during the induction of obsessions. Then after the patients were allowed to act out their compulsions the scales start to lower and when relief is achieved the symptoms have approximately returned to their baseline levels. Repeated measures ANOVA showed a significant symptom x time interaction (p < 0.001). Post-hoc one-way ANOVAs for each symptom revealed that anxiety, agitation, obsessions and compulsions significantly changed over time (p<0.001, Bonferroni corrected), whereas mood and avoidance did not.

### Data acquisition and modelling

LFPs were recorded using the 8180 Sensing Programmer SW (Medtronic Inc.) with a sampling rate of 422Hz. Data were band pass filtered between 3-99 Hz and a band stop filter was used between 47-53 Hz due to a non-physiological artifact appearing in 3 patients. For each 3 minute round in each task the recordings consisted of 30 seconds from each contact pair. The electrode has 4 contacts so all possible combinations result in 6 contact pairs. For the modelling a deep learning architecture called InceptionTime was used, which is the best performing model for time-series classification ^18^. It uses convolutional kernels of different sizes, which are concatenated before feeding them to the next layer. We used samples of 3 second from each hemisphere as input to the network.

### Patient Prediction

We first explored whether the individual patients could be identified using data obtained at rest. Since this experiment was part of a larger study which involved different experiments we used the baselines from the provocation experiment as well as baselines or resting state from other experiments the patients performed. In these other baseline or resting state sessions, LFP recordings were performed when patients sit still for 3 minutes with DBS off either with eyes open or closed. The amount of data available for this task was 10.3 (SD: 4.6) minutes on average. The experiment with least amount of baseline data was used as test set and all others as training set. Overall balanced accuracy for patient prediction was 32.6 % which was significantly above chance level of 9% (p<0.05) as assessed by permutation testing. The patient predictions worked well for patients 2 and 10 with 97% and 98% accuracy respectively, but did not work at all for most patients (see supplementary Figure 2). This indicates that some patients have a highly specific neural signature, whereas others showed comparable signals and that we did not have enough subjects to capture the between subject differences in our model.

### State prediction

Next, we tested whether the model could distinguish between the symptom states across patients. We used data from the symptom provocation experiment. Data for each patient was tested using a model that was trained on data from the other patients in an iterative manner. Overall balanced accuracy was 31% (SD: 6%) while chance level was 25% for the four states. Since this model was computationally expensive to run and the accuracy was within one SD from chance level we did not run permutation testing.

We then tested whether patient-specific models could identify the symptom state a particular patient was in. We therefore trained a model for each patient independently using the first 3 rounds of the data, and tested the state predictions using the last round of the data. For this four-states problem we used balanced accuracy to evaluate the results. However to further gain insight into the performance for each state, sensitivity and specificity were used per state. Since those measures are only defined for binary classification this requires looking at comparisons between each state of interest vs. the rest. That is an unbalanced problem so the sensitivity has a chance level of 25% and the specificity has a chance level of 75%. Further, since sensitivity is arguably of more clinical interest for correctly detecting the state of interest, we focus on that metric. Average balanced accuracy across patients was 38.8% with considerable variation across patients (see Table 3). State prediction was significantly better than chance level in 9 out of 11 patients. Significance was assessed with permutation testing. The models seem to do best for compulsions with an average sensitivity of 52%, followed by obsessions with 41.1%. Sensitivities for baseline and relief are on average 31%. For the patient with the best performing model, patient 2, the results were driven by good performance on baseline and obsessions. For the patients with the next well performing models, patients 4, 5, 8 and 11, the performance was driven by good performance on the compulsions.

**Table 3:**
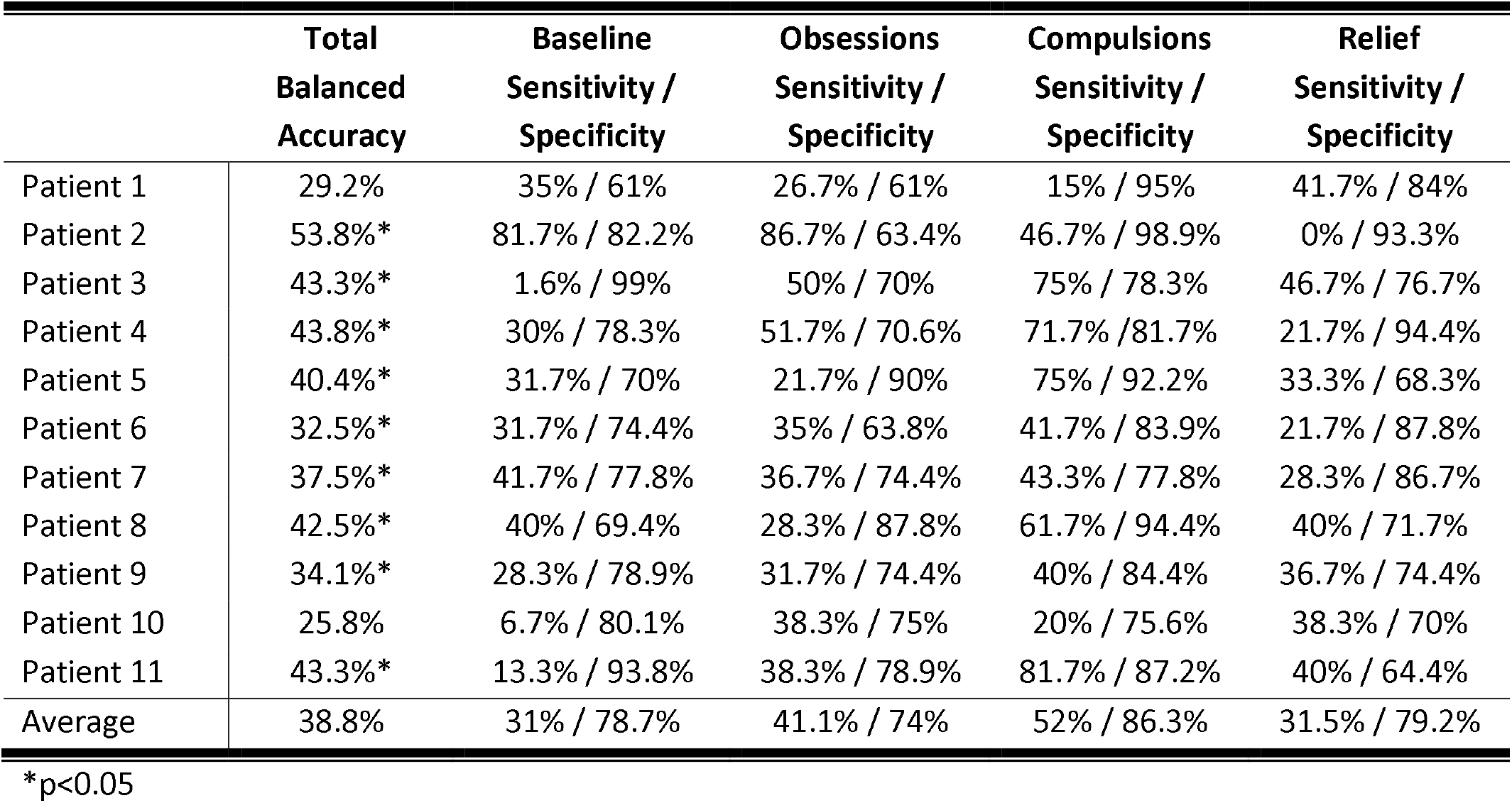
Results from patient level predictions

|  | **Total**<br>**Balanced**<br>**Accuracy** | **Baseline**<br>**Sensitivity /**<br>**Specificity** | **Obsessions**<br>**Sensitivity /**<br>**Specificity** | **Compulsions**<br>**Sensitivity /**<br>**Specificity** | **Relief**<br>**Sensitivity /**<br>**Specificity** |
| --- | --- | --- | --- | --- | --- |
| Patient 1 | 29.2% | 35% / 61% | 26.7% / 61% | 15% / 95% | 41.7% / 84% |
| Patient 2 | 53.8%* | 81.7% / 82.2% | 86.7% / 63.4% | 46.7% / 98.9% | 0% / 93.3% |
| Patient 3 | 43.3%* | 1.6% / 99% | 50% / 70% | 75% / 78.3% | 46.7% / 76.7% |
| Patient 4 | 43.8%* | 30% / 78.3% | 51.7% / 70.6% | 71.7% / 81.7% | 21.7% / 94.4% |
| Patient 5 | 40.4%* | 31.7% / 70% | 21.7% / 90% | 75% / 92.2% | 33.3% / 68.3% |
| Patient 6 | 32.5%* | 31.7% / 74.4% | 35% / 63.8% | 41.7% / 83.9% | 21.7% / 87.8% |
| Patient 7 | 37.5%* | 41.7% / 77.8% | 36.7% / 74.4% | 43.3% / 77.8% | 28.3% / 86.7% |
| Patient 8 | 42.5%* | 40% / 69.4% | 28.3% / 87.8% | 61.7% / 94.4% | 40% / 71.7% |
| Patient 9 | 34.1%* | 28.3% / 78.9% | 31.7% / 74.4% | 40% / 84.4% | 36.7% / 74.4% |
| Patient 10 | 25.8% | 6.7% / 80.1% | 38.3% / 75% | 20% / 75.6% | 38.3% / 70% |
| Patient 11 | 43.3%* | 13.3% / 93.8% | 38.3% / 78.9% | 81.7% / 87.2% | 40% / 64.4% |
| Average | 38.8% | 31% / 78.7% | 41.1% / 74% | 52% / 86.3% | 31.5% / 79.2% |

\*p<0.05

We next tested whether ensemble learning could improve model performance. Five consecutive samples were ensembled together and majority voting was used to determine the prediction for test data. Linked permutations showed that this procedure improved balanced accuracy significantly in 3 patients, decreased significantly in 2 patients, and did not change significantly for the other patients (Figure 3). In patient 2 the accuracy reached 67% and sensitivity for obsessions reached 91.2% while the specificity was similar as before. For compulsions the sensitivity/specificity was 75%/100%. Patient 4 reached accuracy of 54% while specificity/sensitivity for obsessions and compulsions were 75%/72.2% and 91.7%/94.4% respectively.

**Figure 4:**
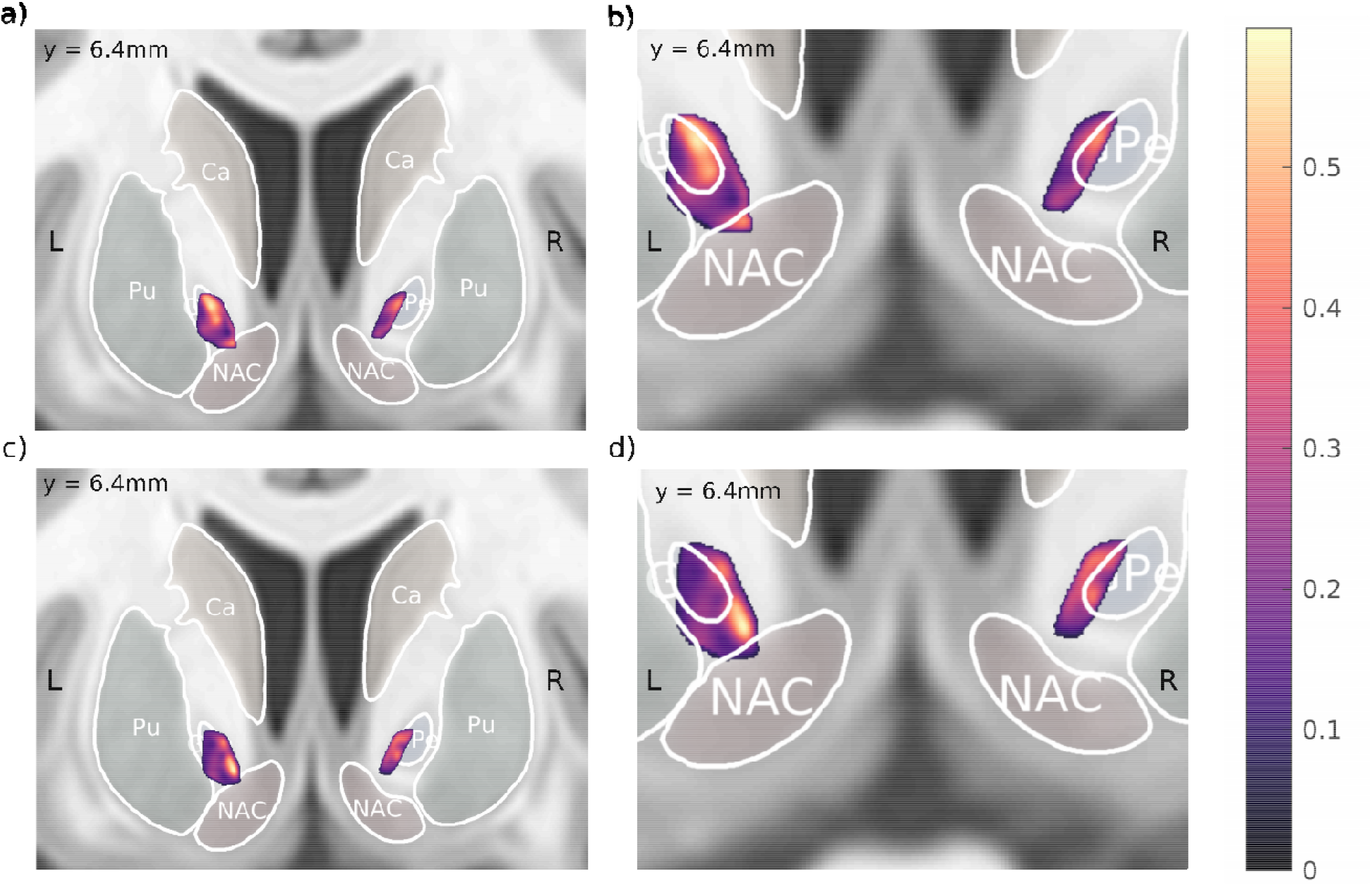
Coronal views of the model performance calculated for obsessions and compulsions. a) Sensitivities for obsessions and b) a zoomed in version with contours of GPe and NAc overlaid. c) Sensitivities for compulsions. d) zoomed in version with contours of GPe and NAc overlaid on sweet spots. The color indicates sensitivity, chance level is 25%. Ca: Caudate, Pu: Putamen, NAC: Nucleus accumbens, GPe: Global pallidus external

### Localization of model performance

To localize the prediction results only adjacent contact pair samples from the test set were fed to the trained model. This was done one hemisphere at a time and only for sensitivity in obsessions and compulsions. Then the resulting 33 sensitivities per hemisphere and state were mapped to the midpoint between the corresponding electrode contacts in MNI space following the approach of Horn et al ^19^. A scattered interpolant was used to fill the spaces between the measurements and then a 0.7mm Gaussian kernel was used to smooth the results to reduce noise.

In Figure 4 the model performance by location for obsessions and compulsions can be seen. For obsessions the highest sensitivities were on the left side and seemed to be higher in gray matter such as the globus pallidus externa (GPe) and the NAc. For compulsions the sensitivities were also higher on the left side and the peak performance was in the white matter just above the NAc, which incidentally was an area of low performance for obsessions.

### Cardiac artifact

In patient 2 there was a heartbeat-like artifact in the left hemisphere in all channel pairs that included contact number 0. This was a known issue in the Activa PC+S because of a fluid leakage past a seal on top of the stimulator which affected contact points 0 and 8 most^20^. The beats per minute were approximately 78-94. Since this artifact could potentially be correlated to symptom states the predictions for patient 2 were repeated with the left artifact free channels copied over the channels with the artifact in the training set and the model re-trained. This had little influence on the accuracies on the test set, with an overall accuracy of 53.8% and state specific sensitivities of 86.7%, 81.7%, 45% and 0% (baselines, obsessions, compulsions and relief).

## Discussion

This study is first of its kind in using deep learning algorithms on deep-brain recordings of psychiatric patients. We aimed to identify a biomarker that is suitable for the use of closed loop DBS in OCD. We induced obsessions and compulsions in eleven OCD patients using a symptom provocation paradigm and trained a deep learning model on time series recorded from the electrodes used during DBS for OCD. Our main finding was that models based on data from multiple patients were hardly better than chance. Models for individual patients were more successful in identifying different symptom states in majority of patients, though with low to moderate accuracy. Therefore, our results suggest that LFP signatures of obsessions and compulsions are highly patient specific.

The performance of our patient-specific models and the identification of compulsive and obsessive states indicates that the model is picking up on an underlying signal of cognitive and/or affective processes. Since obsession and compulsion signals were differentially detected it might mean that there are different underlying signals for these symptom states in the oscillatory activity of the Ventral Striatum. This is further corroborated with the location of model performance map in Figure 4 where the peak in the compulsions on the left side corresponds to low performance for obsessions, which therefore seems to relate to different neural sources. The electrode leads were targeted so that the most ventral contact points were located in the NAc. The other contact points were in the white matter in the vALIC and a part were in the GPe (see Figure 4). All the recordings were in a bipolar configuration where the signal is dominated by neurons local to the electrode and the spatial reach of the LFP signal is in the order of two to five mm, where the distance between the recording contacts increases the spatial reach ^21^. For analyzing performance of the model by location only adjacent contacts were used so the spatial reach was highly localized. The LFP signal reflects postsynaptic mechanisms representing the afferent input activity of the neurons ^22^. Thus, it seems our models are most accurate in predicting obsessions when using signal from gray matter in the GPe and NAc. This signal reflects the input to those regions. Both those regions are part of the cortico-striato-thalamo-cortical (CSTC) loops which are believed to be central to the pathophysiology of OCD^23^. The CSTC receives projections from regions in the frontal cortex which terminate in the striatum and then travel by either a “direct” (net excitatory) or “indirect” (net inhibitory) pathways to the thalamus before returning to the frontal cortex. It has been hypothesized that in OCD the network is biased towards the direct pathway resulting in failure to inhibit abnormal and repetitive behaviors ^24^. The NAc is part of the input to the CSTC network while the GPe is part of the indirect pathway, though recent studies suggest a more complicated role for the GPe where it links information from both direct and indirect pathways^25^. How this fits our results that obsessions are better detected in those regions is not clear although these are close and interconnected structures.

For the compulsions the best accuracy was in the white matter close to the NAc. Traditionally, LFP signals from white matter have been considered to reflect volume conduction from nearby gray matter regions and be otherwise absent. Recently, it was shown that using bipolar or local referencing minimizes the volume conductance effects and there remains some signal in the white matter, which can be correlated to gray matter regions some cm away, thus the LFPs could be from white matter tracts ^26^. The white matter signal is of lower amplitude and spectral power than in gray matter, so that could be one reason obsessions are better distinguished in gray matter. White matter tracts have been increasingly used as targets for DBS in OCD where an increase in response has been associated with targeting the frontopontine tract ^27–29^. But without diffusion MRI data it is hard to find out if the compulsion performance peak overlaps with this tract.

Our result suggest that building patient specific models works better than population models. There is variability between patients in precise target location, clinical presentation and medicine use. As such it should not be surprising that with only 11 patients it is hard to build one model which can predict symptoms for a new patient. Also the prediction of patient identity did not work well. This is probably also because of the variability between patients and may also be partly explained by not having enough data for the model to learn this variability. However, these results also indicate that learning patient specific models may more likely result in clinical tools than population models. But this does mean that in practice once a new patient has been implanted the model first needs to be trained, which has become feasible with the new generation of DBS devices.

This is the largest sample using symptom provocation in OCD while recording LFPs. The symptom provocation tasks were selected before the experiment in consultation with the patients and were highly successful for anxiety, agitation, obsessions and compulsions (Figure 1). In contrast Miller et al^30^ did perioperative symptom provocation in 1 patient while Rappel et al^15^ collected both resting state and symptom provocation LFP data from 2 OCD patients and compared to 4 Parkinson’s disease patients which used the same surgical target as the OCD patients. However, one of the OCD patients dropped out early so they only presented data from the symptom provocation from 1 patient. Miller et al did find gamma oscillations in the NAc modulated by symptom provocation. However they found opposite effects in the two locations measured (2mm apart) suggesting these modulations are variable depending on location. Rappel et al measured LFP in the STN so it is hard to compare that to our data. We are to our knowledge the only group so far presenting results using multi-patient data with symptom provocation.

There are some limitations to this study. Although this is the largest OCD study with sensing LFP data so far, the sample size is small, due to the limited number of investigational devices that were available. Additionally, the recording time was limited, in part due to device limitations on how much data could be continuously recorded before storing it on a computer. This will be better in newer generations of these types of devices. In the patient specific models, we are predicting symptoms in the last round using the other rounds as training data. This means we are predicting symptoms occurring a few minutes later. A more realistic task would be predicting symptoms in a future session, separated by weeks or even months. The model that was used is a black box model, and it is hard to figure out what the model is using to predict the symptom state. Further these are chronically ill patients and as such were on different medications. Due to low sample size we could not control for medication use nor OCD subtypes. In our sample most of the patients were female and eventual non-responders which might have affected the results, though which may be a more important consideration for the group models than the patient-specific models. Due to the nature of the task, where most patients were moving during the compulsion phase (e.g. washing hands), it cannot be ruled out that motion artifacts or motion related changes in neural activity influenced those results. Further experiments will need to take into account and control for this confound which might influence the prediction of compulsions. To analyze model performance by location warping into MNI space was used to localize the electrodes. There is some uncertainty in this, however the algorithm used is specifically optimized for maximal precision of subcortical deformations^31^.

Future work should focus on gathering more data per patient, specifically in a real-life scenario. The individually tailored symptom provocation used here seems to work well to induce symptoms. Newer devices will be capable of storing more data on the device before needing to upload it to a computer. With more data better models could be built and more research done on under what specific conditions the models can detect symptoms. Future research should also work on using explainability to visualize and interpret the model decisions. That would give insight both into how the model identifies symptoms as well as into the disease itself.

## Data Availability

Data available upon reasonable request.

## Acknowledgements

We thank Gaetano Leogrande from Medtronic for his assistance with technical matters. We further thank the students who assisted with the experiment, V.I Visser, S.Y. van Dommelen, N. Runia, M.E. Ottenheim, E.S.A Dijkstra and C.M. de Booi.

## Conflicts of interest

The deep brain stimulation devices were provided by Medtronic for research purposes. The company had no role in the design of the study, data analysis, drafting of the manuscript, or manuscript submission.

## Online Methods

### Participants

Eleven patients with treatment refractory OCD were recruited from the outpatient clinic for DBS at the department of Psychiatry of the Amsterdam University Medical Center, location Academic Medical Center (AMC), in Amsterdam from 2015-2020. Patients were eligible either if they had an existing DBS system implanted and had responded to treatment and needed a stimulator replacement (one patient), or if they were newly indicated for implantation of a DBS system (ten patients). Exclusion criteria was alcohol or substance abuse during last 6 months. The study was approved by the medical ethics board of the AMC and all patients consented to participate in this study and signed an informed consent form. The trial was registered in the Netherlands Trial Register (Trial NL7486).

### Deep brain stimulation

For the implantation two quadripolar electrodes (Model 3389 Medtronics) were used with 4 contact points of 1.5 mm each separated by 0.5 mm. The electrodes were implanted bilaterally through the anterior limb of the internal capsule with the most ventral contact point located in the nucleus accumbens in the plane 3 mm anterior to the anterior commissure (Figure 1). and the 3 upper contact points positioned in the ventral part of the anterior limb of the internal capsule (vALIC) (supplementary Figure 1). The neurostimulator used is the Activa PC + S (Medtronic; model 37604) which allows both for stimulating and recording local field potentials ^8^. The recording and therapy part of the device are completely separated and the therapy part is identical to the Activa PC stimulator from Medtronic.

### Symptom provocation experiment

Two to three weeks after implantation or stimulator replacement the patients underwent a symptom provocation task before. For the newly indicated patients this was before the DBS device was activated for the first time with the exception of patient 2 which had his experiment several weeks later than usual. For the patients using DBS the device was switched off during the experiment. First the symptom severity was assessed using the Yale-Brown Obsessive Compulsive Scale (Y-BOCS) ^32^, the Hamilton Depression Rating Scale (HAM-D) ^33^ and the Hamilton Anxiety Rating Scale (HAM-A)^34^ for OCD, depressed mood and anxiety respectively. Then the patient underwent 4 rounds of a symptom provocation sequence (depicted in Figure 2). First the patient was instructed to sit calmly and watch a neutral movie for three minutes while LFPs were recorded. Movie watching was chosen to minimize obsessions. Then obsessions were induced in a patient specific manner (see Table 2). For example patients with contamination obsessions were asked to touch the floor. Once obsessions had been induced the patient was instructed to sit and focus on their obsessions for three minutes while LFPs were recorded. Then three minutes of recording followed where the patient was allowed to act out their compulsions, e.g. washing hands for contamination obsessions. After that, the patient could continue to act out the compulsions if needed until feeling relieved. Then the patient was instructed to sit calmly while LFPs were recorded. Recordings were done in left and right hemisphere simultaneously. Before and after each task the patient rated their symptoms on a visual analogue scale (VAS) measuring anxiety, agitation, mood, obsessions, compulsions and avoidance.

The visual analog scales were entered into a repeated measure ANOVA to test for interaction of symptoms (6 items) and time (7 VAS measurements). If an interaction was significant, post-hoc tests were carried out

### Data acquisition and modelling

LFPs were recorded using the 8180 Sensing Programmer SW (Medtronic Inc.) with a sampling rate of 422Hz using hardware 0.5 Hz high pass filtering and 100 Hz low pass filtering ^8^. Due to amplifier settling the first 1000 samples were removed from each 30 second recording. Furthermore because of the presence of a marker signal at 104.5Hz to measure channel saturation ^20,35^, the signal was band passed filtered between 3-99 Hz using a zero phase finite impulse response filter using MNE-python ^36^. Three Hz was chosen as a high pass threshold to minimize artifacts. Further there were non-physiological artifacts in the frequency bands between 47-53 Hz in 3 patients so a band stop filter was used to filter those bands out in all patients.

Since this experiment was part of a larger study there was baseline or resting state data available from several other experiments. The baselines from this experiment as well as baselines from other sessions in other experiments the patient performed were used for patient prediction. These other baseline data consisted of resting state data where the patient sat still for 3 minutes with eyes either open or closed. The amount of baseline data available on average per patient was 10.3 minutes (SD: 4.6). The baselines were split into training and test set by using the experiment with smallest amount of data as test and all other sessions as training. The training set was then sampled randomly 300 times and the test set 100 times per patient. The duration of the samples was three seconds and was chosen using the validation set performance. Twenty percent of the training set was used as a validation set for model selection. For the patient predictions, only data from the outermost contact pair were used since most baseline data was available of that channel pair for each patient.

For group level state prediction, the data from the described experiment were used. One patient was selected randomly as the test patient and one patient as validation set. The rest were used for training. This was then repeated for all sets of test patients. We used 180 random three second samples per round per state, in total 2880 samples per patient for the training set. We used oversampling by a factor of 3 for the training set as a form of data augmentation. For the test set 60 samples per round per state were used or 240 samples per patient. For single patient state prediction, the first three rounds in the experiment were used for training and the last round for testing. Twenty percent of the training set was used as validation set. The number and size of samples was the same as for the group level prediction. In all experiments both data from the left and right electrode were used. Each three-minute task in each round consisted of 30 seconds with measurement from each channel from the electrode. There were 4 contact points and thus 6 independent bipolar channels measured consisting of slightly different positions in the patient’s brain. Each channel measures the voltage between two contacts on the electrode.

The model used was InceptionTime ^18^ which was recently shown to have state of the art performance on time-series classification. The model consists of inception blocks and residual blocks. For the inception blocks, convolutional filters of three different sizes and a max pooling filter are simultaneously slid over the input. The results from these operations are then concatenated to form the input to the next block. There are three inception blocks in one residual block. The input of each residual block is added to the output, forming a residual connection. The number of residual blocks, sizes of the kernels and number of filters for the convolutional layers were found using hyper-parameter tuning on the validation set. After the residual blocks a global average pooling layer was used and then a fully connected layer with the number of neurons the same as the number of classes being predicted.

The model was trained in PyTorch using a learning rate of 1e-4 and a batch size of 128 using the Adam optimizer ^37^. If the validation loss stopped decreasing for three epochs the learning rate was reduced tenfold and if the validation loss stopped decreasing for six epochs the training was stopped. As in Fawaz et al^18^, for every task few trained five instances of the model with different random initializations and the outputs averaged for the final predictions. This is in line with Mehrer et al^38^, who show that predictions are more stable when using ensembles of deep learning models. Permutation testing was used to assess significance with 120 permutations per model ensemble. To further test the capabilities of the patient specific models consecutive 5 samples were drawn from the test set. The total amount of samples drawn was reduced by a factor of 5 so it was the same in seconds as before. Then the fitted model was used to predict states for the consecutive samples. Then majority voting among the 5 samples was used to make a prediction for the total period. To see if this method was more accurate than using a single sample, linked permutations were used where the labels were permuted and both methods used on the permuted labels and a null distribution of the difference between the methods was built.

To analyze differences in performance by location the approach of Horn et al^19^ was used. A pre-surgery anatomical T1 sequence of an MRI-scan was linearly coregistered to a post-surgery CT scan using Advanced Normalization Tools (ANTs) ^39^. These were then normalized into the ICBM 2009b NLIN asymmetric MNI space ^40^ using ANTs. Then the electrode trajectories and contact positions were reconstructed using the PaCER method ^41^. The atlas used for visualization was from Pauli et al^42^. The model sensitivity for obsessions and compulsions was calculated per hemisphere for channels using measurements from adjacent contact points on the test set. For four contact points this results in 3 measurements per patient per hemisphere. Then each measurement was mapped to the midpoint between the electrode contacts in the common space. The measurements were then interpolated using a scattered interpolant and the result smoothed with a Gaussian kernel of 0.7 mm. The resulting sweet spot map was then visualized using Lead Group ^43^.

## Footnotes

a https://www.medtronic.com/us-en/healthcare-professionals/products/neurological/deep-brain-stimulation-systems/percept-pc.html

## Notes

### Clinical Trial

NL7486

### Funding Statement

No external funding.

### Author Declarations

The study has been reviewed and approved by the Institutional Review Board of the Academic Medical Center Amsterdam.

## References

1. Denys, D. Pharmacotherapy of Obsessive-compulsive Disorder and Obsessive-Compulsive Spectrum Disorders. Psychiatric Clinics of North America 29, 553–584 (2006).

2. Alonso, P. et al. Deep Brain Stimulation for Obsessive-Compulsive Disorder: A Meta-Analysis of Treatment Outcome and Predictors of Response. PLoS One 10, e0133591 (2015).

3. Denys, D. et al. Efficacy of Deep Brain Stimulation of the Ventral Anterior Limb of the Internal Capsule for Refractory Obsessive-Compulsive Disorder: A Clinical Cohort of 70 Patients. Am. J. Psychiatry (2020). doi:10.1176/appi.ajp.2019.19060656

4. Graat, I. et al. Long-term Outcome of Deep Brain Stimulation of the Ventral Part of the Anterior Limb of the Internal Capsule in a Cohort of 50 Patients With Treatment-Refractory Obsessive-Compulsive Disorder. Biol. Psychiatry (2020). doi:10.1016/j.biopsych.2020.08.018

5. Duarte, G. S. & do Couto, F. S. Efficacy, effect on mood symptoms, and safety of deep brain stimulation in refractory obsessive-compulsive disorder: A systematic review and meta-analysis. Journal of Clinical Psychiatry (2020). doi:10.4088/JCP.19r12821

6. de Koning, P. P., Figee, M., van den Munckhof, P., Schuurman, P. R. & Denys, D. Current Status of Deep Brain Stimulation for Obsessive-Compulsive Disorder: A Clinical Review of Different Targets. Curr. Psychiatry Rep. 13, 274–282 (2011).

7. Coenen, V. A. et al. The medial forebrain bundle as a target for deep brain stimulation for obsessive-compulsive disorder. CNS Spectr. 22, 282–289 (2017).

8. Stanslaski, S. et al. Design and validation of a fully implantable, chronic, closed-loop neuromodulation device with concurrent sensing and stimulation. IEEE Trans. Neural Syst. Rehabil. Eng. 20, 410–421 (2012).

9. Bouthour, W. et al. Biomarkers for closed-loop deep brain stimulation in Parkinson disease and beyond. Nat. Rev. Neurol. 15, 343–352 (2019).

10. Tinkhauser, G. et al. Directional local field potentials: A tool to optimize deep brain stimulation. Mov. Disord. 33, 159–164 (2018).

11. Kim, S. et al. Closed-Loop Neuromodulation for Parkinsons Disease: Current State and Future Directions. IEEE Trans. Mol. Biol. Multi-Scale Commun. (2020). doi:10.1109/TMBMC.2020.3036756

12. Houston, B., Thompson, M., Ko, A. & Chizeck, H. A machine-learning approach to volitional control of a closed-loop deep brain stimulation system. J. Neural Eng. 16, 016004 (2019).

13. Opri, E. et al. Chronic embedded cortico-thalamic closed-loop deep brain stimulation for the treatment of essential tremor. Sci. Transl. Med. 12, eaay7680 (2020).

14. Tastevin, M., Spatola, G., Régis, J., Lançon, C. & Richieri, R. Deep brain stimulation in the treatment of obsessive-compulsive disorder: current perspectives. Neuropsychiatr. Dis. Treat. 15, 1259–1272 (2019).

15. Rappel, P. et al. Subthalamic theta activity: a novel human subcortical biomarker for obsessive compulsive disorder. Transl. Psychiatry 8, 118 (2018).

16. Olsen, S. T. et al. Case Report of Dual-Site Neurostimulation and Chronic Recording of Cortico-Striatal Circuitry in a Patient With Treatment Refractory Obsessive Compulsive Disorder. Front. Hum. Neurosci. 14, 423 (2020).

17. LeCun, Y., Bengio, Y. & Hinton, G. Deep learning. Nature 521, 436–444 (2015).

18. Ismail Fawaz, H. et al. InceptionTime: Finding AlexNet for time series classification. Data Min. Knowl. Discov. 34, 1936–1962 (2020).

19. Horn, A., Neumann, W. J., Degen, K., Schneider, G. H. & Kühn, A. A. Toward an electrophysiological “Sweet spot” for deep brain stimulation in the subthalamic nucleus. Hum. Brain Mapp. (2017). doi:10.1002/hbm.23594

20. Quinn, E. J. et al. Beta oscillations in freely moving Parkinson’s subjects are attenuated during deep brain stimulation. Mov. Disord. (2015). doi:10.1002/mds.26376

21. Lempka, S. F. & McIntyre, C. C. Theoretical Analysis of the Local Field Potential in Deep Brain Stimulation Applications. PLoS One (2013). doi:10.1371/journal.pone.0059839

22. Buzsáki, G., Anastassiou, C. A. & Koch, C. The origin of extracellular fields and currents-EEG, ECoG, LFP and spikes. Nature Reviews Neuroscience (2012). doi:10.1038/nrn3241

23. Milad, M. R. & Rauch, S. L. Obsessive-compulsive disorder: Beyond segregated cortico-striatal pathways. Trends in Cognitive Sciences 16, 43–51 (2012).

24. Ahmari, S. E. & Dougherty, D. D. DISSECTING OCD CIRCUITS: From ANIMAL MODELS to TARGETED TREATMENTS. Depression and Anxiety (2015). doi:10.1002/da.22367

25. Okamoto, S. et al. Overlapping Projections of Neighboring Direct and Indirect Pathway Neostriatal Neurons to Globus Pallidus External Segment. iScience (2020). doi:10.1016/j.isci.2020.101409

26. Mercier, M. R. et al. Evaluation of cortical local field potential diffusion in stereotactic electro-encephalography recordings: A glimpse on white matter signal. Neuroimage (2017). doi:10.1016/j.neuroimage.2016.08.037

27. Liebrand, L. C. et al. Individual white matter bundle trajectories are associated with deep brain stimulation response in obsessive-compulsive disorder. Brain Stimul. (2019). doi:10.1016/j.brs.2018.11.014

28. Li, N. et al. A unified connectomic target for deep brain stimulation in obsessive-compulsive disorder. Nat. Commun. (2020). doi:10.1038/s41467-020-16734-3

29. Haber, S. N., Yendiki, A. & Jbabdi, S. Four Deep Brain Stimulation Targets for Obsessive-Compulsive Disorder: Are They Different? Biological Psychiatry (2020). doi:10.1016/j.biopsych.2020.06.031

30. Miller, K. J., Prieto, T., Williams, N. R. & Halpern, C. H. Case Studies in Neuroscience: The electrophysiology of a human obsession in nucleus accumbens. J. Neurophysiol. 121, 2336–2340 (2019).

31. Ewert, S. et al. Optimization and comparative evaluation of nonlinear deformation algorithms for atlas-based segmentation of DBS target nuclei. Neuroimage (2019). doi:10.1016/j.neuroimage.2018.09.061

32. Goodman, W. K. et al. The Yale-Brown Obsessive Compulsive Scale. II. Validity. Arch. Gen. Psychiatry 46, 1012–6 (1989).

33. Hamilton, M. A Rating Scale for Depression. J. Neurol. Neurosurg. Psychiat 23, 56–62 (1960).

34. Hamilton, M. Hamilton Anxiety Rating Scale (HAM-A). J. Med. 61, 81–82 (1959).

35. Herron, J. et al. Bi-directional brain interfacing instrumentation. in I2MTC 2018 - 2018 IEEE International Instrumentation and Measurement Technology Conference: Discovering New Horizons in Instrumentation and Measurement, Proceedings (2018). doi:10.1109/I2MTC.2018.8409795

36. Gramfort, A. et al. MEG and EEG data analysis with MNE-Python. Front. Neurosci. 7, 267 (2013).

37. Kingma, D. P. & Ba, J. L. Adam: A method for stochastic optimization. in 3rd International Conference on Learning Representations, ICLR 2015 - Conference Track Proceedings (2015).

38. Mehrer, J., Spoerer, C. J., Kriegeskorte, N. & Kietzmann, T. C. Individual differences among deep neural network models. Nat. Commun. 11, 5725 (2020).

39. Avants, B. B., Epstein, C. L., Grossman, M. & Gee, J. C. Symmetric diffeomorphic image registration with cross-correlation: Evaluating automated labeling of elderly and neurodegenerative brain. Med. Image Anal. 12, 26–41 (2008).

40. Fonov, V., Evans, A., McKinstry, R., Almli, C. & Collins, D. Unbiased nonlinear average age-appropriate brain templates from birth to adulthood. Neuroimage (2009). doi:10.1016/s1053-8119(09)70884-5

41. Husch, A. V. Petersen, M., Gemmar, P., Goncalves, J. & Hertel, F. PaCER - A fully automated method for electrode trajectory and contact reconstruction in deep brain stimulation. NeuroImage Clin. (2018). doi:10.1016/j.nicl.2017.10.004

42. Pauli, W. M., Nili, A. N. & Tyszka, J. M. A high-resolution probabilistic in vivo atlas of human subcortical brain nuclei. Sci. Data 2018 5 5, 180063 (2018).

43. Treu, S. et al. Deep brain stimulation: Imaging on a group level. Neuroimage (2020). doi:10.1016/j.neuroimage.2020.117018

