## Supplemental Figures for "The ventral striatum harbours patient specific intracranial neural signatures of obsessions and compulsions"

Supplementary material

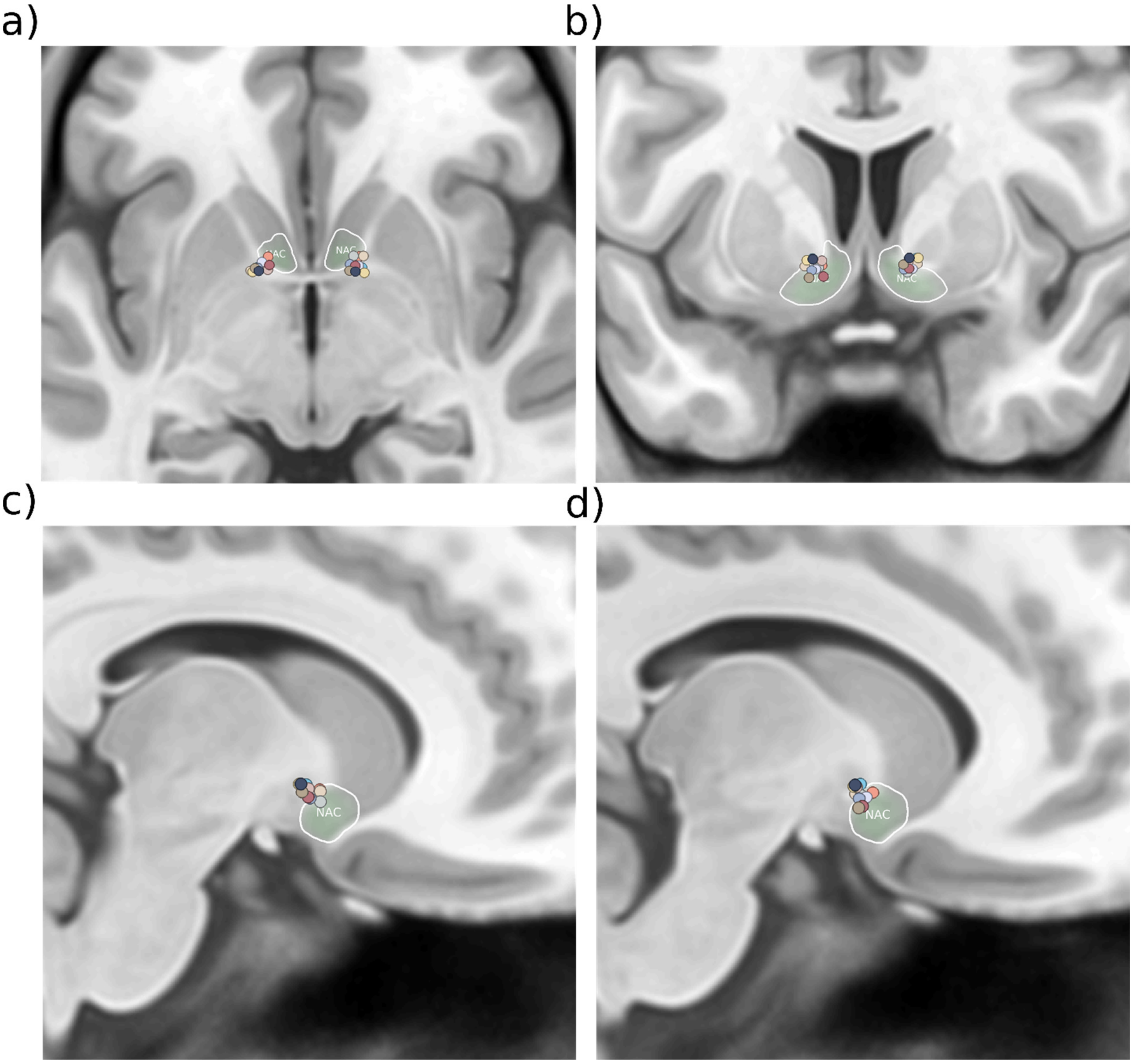


Supplementary Figure 1: Location of the most ventral contact on each electrode for each patient. a) Axial view b) coronal view, c) sagittal view of right targets and d) sagittal view of left targets. Positions transformed and co-registered in MNI space.

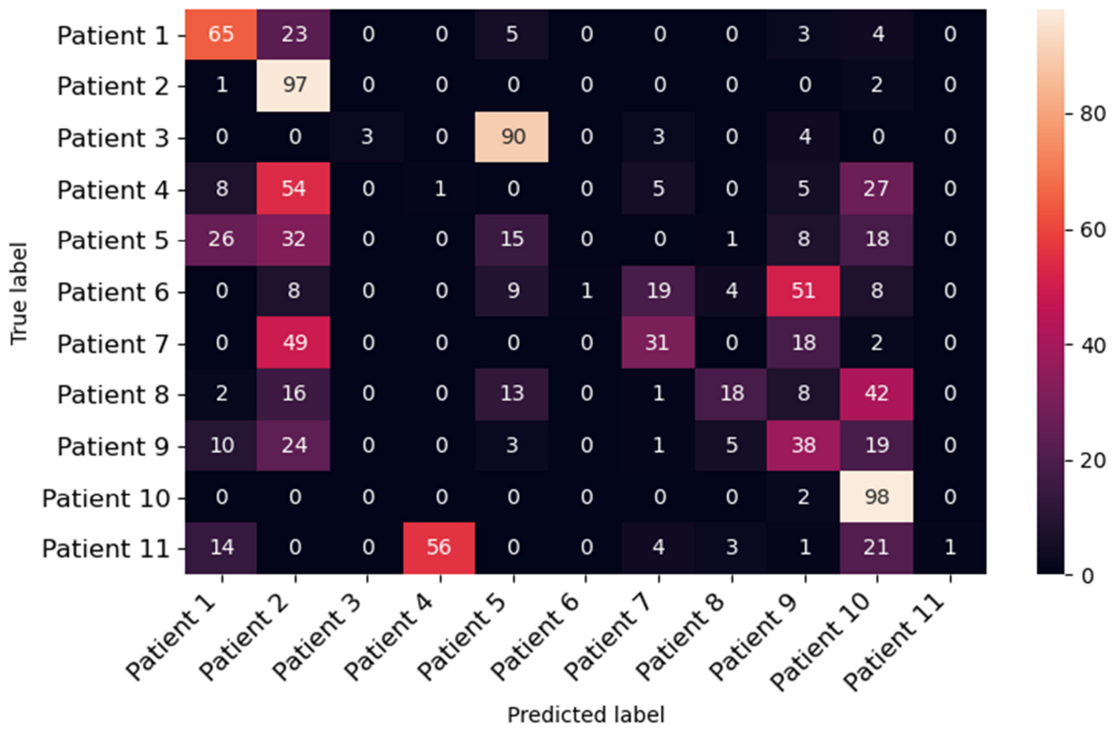


Supplementary Figure 2: Confusion matrix for patient prediction. Each row shows the true labels while the columns show the predicted labels. The diagonal indicates correct predictions.
